# Antibody responses to SARS-CoV-2 in COVID-19 patients: the perspective application of serological tests in clinical practice

**DOI:** 10.1101/2020.03.18.20038018

**Authors:** Quan-xin Long, Hai-jun Deng, Juan Chen, Jie-li Hu, Bei-zhong Liu, Pu Liao, Yong Lin, Li-hua Yu, Zhan Mo, Yin-yin Xu, Fang Gong, Gui-cheng Wu, Xian-xiang Zhang, Yao-kai Chen, Zhi-jie Li, Kun Wang, Xiao-li Zhang, Wen-guang Tian, Chang-chun Niu, Qing-jun Yang, Jiang-lin Xiang, Hong-xin Du, Hua-wen Liu, Chun-hui Lang, Xiao-He Luo, Shao-bo Wu, Xiao-ping Cui, Zheng Zhou, Jing Wang, Cheng-jun Xue, Xiao-feng Li, Li Wang, Xiao-jun Tang, Yong Zhang, Jing-fu Qiu, Xia-mao Liu, Jin-jing Li, De-chun Zhang, Fan Zhang, Xue-fei Cai, De-qiang Wang, Yuan Hu, Ji-hua Ren, Ni Tang, Ping Liu, Qin Li, Ai-long Huang

## Abstract

**Background:** We aim to investigate the profile of acute antibody response in COVID-19 patients, and provide proposals for the usage of antibody test in clinical practice.

**Methods:** A multi-center cross-section study (285 patients) and a single-center follow-up study (63 patients) were performed to investigate the feature of acute antibody response to SARS-CoV-2. A cohort of 52 COVID-19 suspects and 64 close contacts were enrolled to evaluate the potentiality of the antibody test.

**Results:** The positive rate for IgG reached 100% around 20 days after symptoms onset. The median day of seroconversion for both lgG and IgM was 13 days after symptoms onset. Seroconversion of IgM occurred at the same time, or earlier, or later than that of IgG. IgG levels in 100% patients (19/19) entered a platform within 6 days after seroconversion. The criteria of ‘IgG seroconversion’ and ‘> 4-fold increase in the IgG titers in sequential samples’ together diagnosed 82.9% (34/41) of the patients. Antibody test aided to confirm 4 patients with COVID-19 from 52 suspects who failed to be confirmed by RT-PCR and 7 patients from 148 close contacts with negative RT-PCR.

**Conclusion:** IgM and IgG should be detected simultaneously at the early phase of infection. The serological diagnosis criterion of seroconversion or the ‘>; 4-fold increase in the IgG titer’ is suitable for a majority of COVID-19 patients. Serologic test is helpful for the diagnosis of SARS-CoV-2 infection in suspects and close contacts.

## Introduction

The rapid spread of Coronavirus Disease 2019 (COVID-19) worldwide has raised concern around the world. The outbreak of COVID-19 first started in Wuhan of China. With a dramatic increase in daily confirmed global cases, the World Health Organization has declared as a global pandemic on March 12, 2020. As of March 7, 2020, a total of 80813 COVID-19 cases have been confirmed in China and 21110 confirmed cases have been reported in 93 countries/ territories/ areas outside of China.

Early case detection is one of the most important public health interventions in controlling the spread of COVID-19. COVID-19 cases can be identified based on exposure status, symptoms and chest imaging, but the confirmation of infection requires nucleic acid testing of nasal, pharyngeal or anal swab. Although real-time reverse transcription PCR (RT-PCR)-based viral RNA detection is the sensitive and accurate way to confirm the diagnosis of SARS-CoV-2 infection in practice, dozens of suspects with clinical symptoms were failed to be diagnosed by RT-PCR test. The risk of falsely-negative results may be introduced by RT-PCR method due to several possible factors, such as quality of specimen collection, PCR reagents from different sources, multi-steps of RNA preparation and fluctuations of virus load in different phases of SARS-CoV-2 infection. Due to the limitations of RT-PCR, serum specific antibody detection for COVID-19 is advisable because of the relatively short time to diagnosis and the ability to probe an active immune response against the virus.

Human immune responses to a novel pathogen with both innate and adaptive arms. One aspect of the adaptive immunity is humoral response that features the production of antibodies recognizing specific determinants of antigens called epitopes. Some of the antibodies produced can protect the host from future infection by the same pathogens. Studies on severe acute respiratory syndrome (SARS) and Middle East respiratory syndrome (MERS) showed that antibodies were detectable in 80%-100% patients at 2 weeks after illness onset [1, 2, 3, 4], and the antibodies can persist for at least 12 years [5]. Moreover, the profile of antibody responses might be correlated to the severity of disease and outcome in MERS [6, 7]. Currently, as antibody detection for COVID-19 has not been broadly developed, the antibody response against SARS-CoV-2 remains nearly unknown. In this study, we depicted the profiles of acute antibody response against SARS-CoV-2 through a cross-section analysis of 285 patients and a follow-up study of 63 patients. We also demonstrated the clinical application of antibody test in facilitating the diagnosis of COVID-19 both in suspects and asymptomatic close contacts.

## Methods

### Patients

A total of 285 COVID-19 patients for the cross-section study were enrolled from 3 designated hospitals of Chongqing, a province-level municipality adjacent to Hubei province,which is the start-point and at the center of COVID-19 outbreak in China. These three hospitals, Chongqing Three Gorges Central Hospital (TGH), Yongchuan Hospital Affiliated to Chongqing Medical University (CQMU) (YCH), and The Public Health Center of Chongqing (PHCC), were assigned by the Chongqingmn municipal people’s government to admit patients from 3 designated areas indicated in Fig. S1A. All the patients enrolled were confirmed to be infected with SARS-CoV-2 by RT-PCR assay for nasal and pharyngeal swab specimens. For the follow-up cohort, serum samples of 63 patients from YCH were taken at 3-days interval from Feb 8, 2020 to hospital discharge (supplementary figure1B). To evaluate the potentiality of the serological test, a cohort of 52 COVID-19 suspects who failed to be confirmed by RT-PCR were enrolled from Wanzhou People’s Hospital. A serological survey for asymptomatic SARS-CoV-2 infection were conducted in a cohort of close contacts containing 164 persons, which was identified by Chongqing CDC in Wanzhou, Chongqing. The study was approved by the Ethics Commission of Chongqing Medical University (KY-2020-03). Written informed consent was waived by the Ethics Commission of the designated hospital for emerging infectious diseases.

### Detection of the IgG and IgM against SARS-CoV-2

To measure the level of IgG and IgM against SARS-CoV-2, serum samples were collected from the patients. All the serum samples were inactivated at 56 °C for 30 min and stored at -20 °C before testing. The IgG and IgM antibody against SARS-CoV-2 in plasma samples were tested using Magnetic Chemiluminescence Enzyme Immunoassay (MCLIA) kit supplied by Bioscience (Chongqing) Co., Ltd, China (CFDA approved), according to the manufacturer’s instructions. The MCLIA for IgG or IgM detection was developed based on double-antibodies sandwich immunoassay. The recombinant antigens containing the nucleoprotein and a peptide from spike protein of SARS-CoV-2 were conjugated with fluorescein isothiocyanate (FITC) and immobilized on the anti-FITC antibody conjugated magnetic particles. Alkaline phosphatase conjugated human IgG/IgM antibody was used as the detection antibody. The tests were conducted on an automated magnetic chemiluminescence analyzer (Axceed 260, Bioscience, China) according to the manufacturer’s instructions. All the tests were performed under strict biosafe conditions. Antibody levels were expressed by the chemiluminescence signal compared to the cutoff value(S/CO).

### Statistical analysis

Continuous variables were expressed as median (interquartile range, IQR) and compared with the Mann-Whitney U test; categorical variables were expressed as number (%) and compared by χ^2^ test or Fisher’s exact test. P value less than 0.05 was considered statistically significant. Statistical analyses were performed using the *R* software, version 3.6.0.

## Results

### The profile of antibody responses in 285 cases of COVID-19 patients

285 cases of COVID-19 patients admitted to 3 designated hospitals were enrolled in this cross-section study. The median age of these enrolled patients was 47 years (IQR, 34-56 years) and 55.4% were males. 103 patients had an history of exposure to transmission sources, while 262 patients with clear records of symptoms onset. 39 of 285 cases were classified as severe or critical illness condition according to the guidelines (Table S1). For laboratory parameters of the patients, white blood cell, neutrophil D-dimer, hypersensitive troponin I, procalcitonin, CRP (C-reactive protein), lactate dehydrogenase, alanine aminotransferase and aspartate aminotransferase were higher in severe/critical patients, while mild patients had higher level of lymphocyte counts, which is consistent with recent reports [8, 9] (table S2).

Antibody detection rate based on the number of days after onset of symptoms were determined and summarized in Figure 1A. 363 serum samples from 262 patients with clear records of symptoms onset were included in this analysis. The positive rates of IgG and IgM antibodies increased gradually along with days after symptoms onset. The positive rate of IgG reached 100% at around 17-19 days after symptoms onset, while IgM seroconversion rate reached its peak of 94.1% at around 20-22 days after symptoms onset (Fig.1A). 262 patients were further divided into 4 groups based on symptoms onset days(1 week, 2 weeks, 3 weeks and > 3 weeks of symptoms onset). During the first 3 weeks of symptoms onset, there was an increase in the titer of IgG and IgM antibodies to SARS-CoV-2 (Fig.1B). However, the antibody level IgM showed a slightly decrease in the patient group with > 3 weeks of symptoms onset when compared with the > 3 weeks group (Fig.1B). The antibody level of the patients in different clinical courses (Non-severe *vs*. severe) were also compared. There were 20 severe patients who were at the 2^nd^ week after symptoms onset when sampling, and 13 severe patients were at the 3^rd^ week when sampling. As shown in Fig. 1C, IgG and IgM titers in severe group were higher than those in the non-severe group, although significant statistic difference is only observed in IgG level of 2 weeks group (P=0.001).

**Figure 1.**
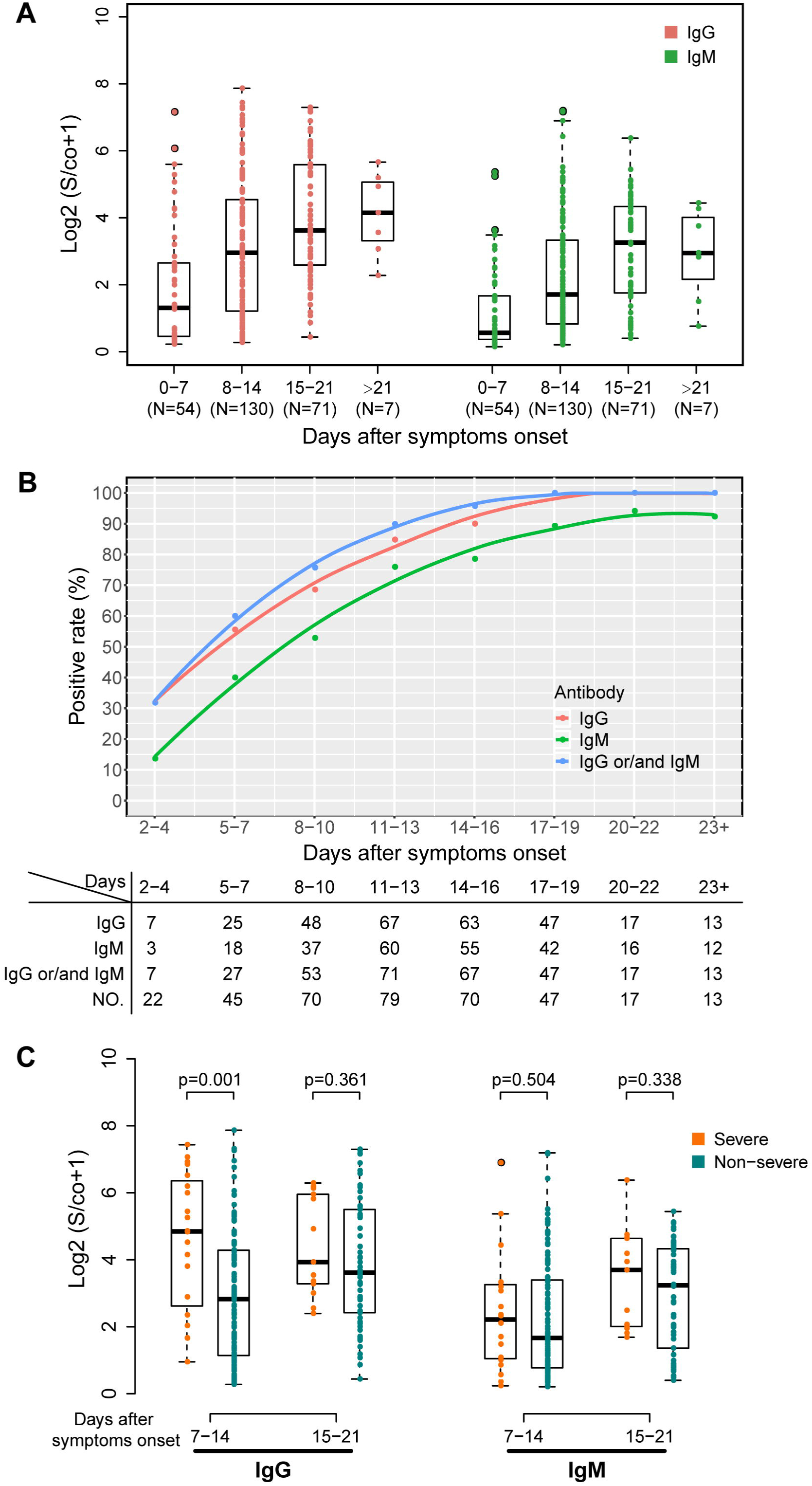
Antibody response against SARS-CoV-2 among COVID-19 patients from cross-section cohort. A. Antibody positive rate based on days after symptoms onset. Patients from cross-section cohort were grouped based on their symptoms onset days (2-4 days, 5-7 days, 8-10 days, 11-13 days, 14-16 days, 17-10 days, 20-22 days and >23 days), antibody positive rates were calculated in different subgroup and present together. B. The relative quantitative titer of antibodies against SARS-CoV-2 in patients with different symptom onset. C. The comparison of relative quantitative titer of antibodies against SARS-CoV-2 between severe and non-severe patients.

### Seroconversion time of the IgG and IgM in COVID-19 patients

To investigate the longitudinal profiles in the IgG and IgM, 63 confirmed COVID-19 patients in the cross-section study were followed up further. A total of 281 sequential serum samples were collected and tested in parallel for the IgG and IgM against SARS-CoV-2. Of the 63 patients, the overall seroconversion rate was 96.8% (61/63) over the follow-up period. Two patients, a 11-year old girl and her mother, maintained IgG and IgM-negative status during hospitalization, indicating that they did not undergo serological conversion during the observation period. Unfortunately, these two patients were lost for follow-up and the delayed seroconversion cannot be excluded. Serological courses could be followed for 27 patients who were initially seronegative and then underwent a seroconversion during the observation (IgG seroconversion in 19 patients, IgM seroconversion in 20 patients and both seroconversion in 13 patients). All these patients achieved a seroconversion of IgG or IgM within 20 days after symptoms onset. The median day of seroconversion for both lgG and IgM was 13 days (after symptoms onset) (Fig. 3A). Three types of seroconversion can be observed: synchronous seroconversion of IgG and IgM (10 cases); IgM seroconverted earlier than that of IgG (7 cases); IgM seroconverted later than that of IgG (10 cases). The longitudinal changes of antibody in 6 representative patients of three types were shown in Fig. 3B-D and Fig. S2. Based on this finding, we recommend that either IgM or IgG seroconversion be used as a confirmation criterion of recent SARS-CoV-2 infection. We tried to identify the factors associated with the different seroconversion types, but found no association between these types and age or gender or hospitalization time (data not shown).

**Fig. 2.**
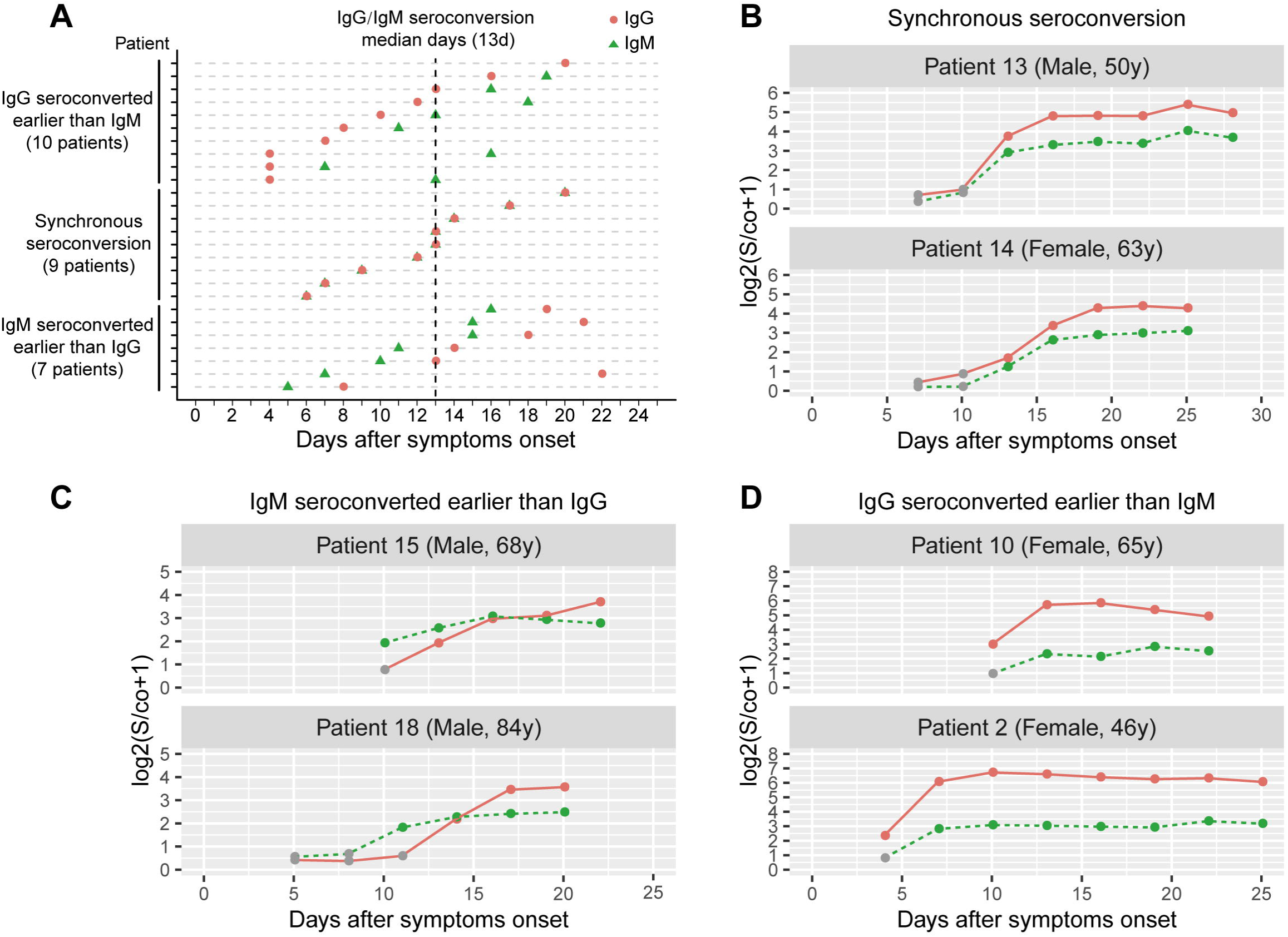
Seroconversion time of the antibodies against SARS-CoV-2 in COVID-19 patients. A. Seroconversion time of the antibodies. Among the 63 patients, 26 achieved seroconversions in the observation. The seroconversion points of each patient were plotted against the days after symptoms onset. The red dots and green triangles indicate the seroconversion points of IgG and IgM respectively. IgM seroconversions of 7 patients were earlier than their IgG seroconversion. IgG seroconversions were earlier in 10 patients, and 9 patients had the IgG and IgM seroconversion almost simultaneously. Median days of both of the seroconversions are 13 days. **B**-**D** showed typical examples of the 3 seroconversion types.

**Fig. 3.**
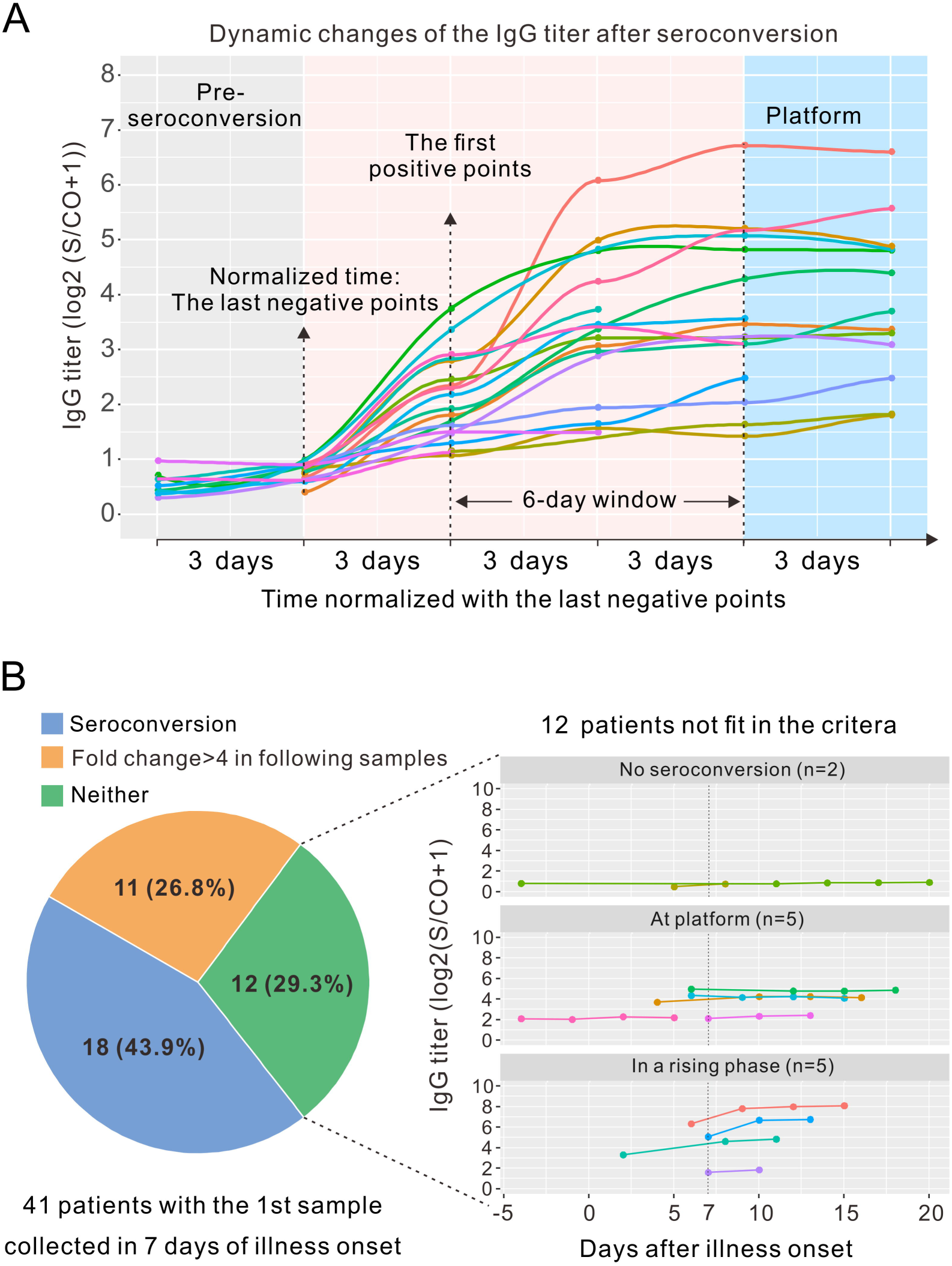
Dynamic change of the IgG antibody during acute response. A. Time course of the IgG against SARS-CoV-2. A relatively complete time course of the IgG response was observed in 19 patients. The last negative time points were normalized. All the IgG titers entered a plateau within 6 days after the first positive samples and the IgG titers were largely different at the platform for different patients. **B. Diagnosis efficacy of the serological test criteria recommended for MERS**. Either seroconversion or a 4-fold increase in the IgG titer in acute (ideally during the first week of illness) and convalescent serum samples was recommended by the WHO as confirmation criteria. We tested whether these criteria are suitable for COVID-19. Among the 41 patients, 29 can be diagnosed by these criteria and 5 cannot.

### Dynamic changes in the titer of the IgG in COVID-19 patients

To analyzed the dynamics changes of the IgG levels in acute response, we normalized the time of the last negative test points of 19 patients who underwent IgG seroconversion during hospitalization. As shown in Fig. 3A, the IgG levels in all the patients reached the platform in 6 days after the first positive points (seroconversion time). However, the IgG levels at the platform in different patients varied largely (more than 20-fold). The IgM titers also showed a similar profile of dynamic change (Fig. S3).

As recommended by the WHO, a seroconversion or a 4-fold increase in the IgG titer in acute (ideally during the first week of illness) and convalescent serum samples can be used to confirm MERS-CoV infection. We thus analyzed if this criterion is also suitable for the diagnosis of COVID-19 [10]. There were 41 patients who were sampled during the first week of illness. As shown in Fig. 3B, 18 of the patients (43.9%) who were IgG negative at the first week seroconverted afterwards (in the observation period). Eleven patients (26.8%) were IgG positive for the first samples, and they eventually achieved a ≥ 4-fold increase in the IgG titer at some points later. However, other 12 patients fit none of the 2 criteria. Among the 12 cases, 2 of them did not seroconvert during the hospitalization. Five of them seemed to be in a rising phase of IgG and might achieve a ≥ 4-fold increase in the future. However, 5 of the patients seemed to enter a plateau of the IgG titer at their first samples (Fig. 3B, lower panel). It is very unlikely that their IgG titer would increase > 4-fold sometime in the near future. Overall, 82.9% (34/41) of the patients fit in the 2 serological criteria above, and 12.2% (5/41) patients already entered the platform of IgG titer at the time of the first sampling (even though in 7 days of illness onset). Very few patients (2/41, 4.9%) did not achieve a seroconversion before discharge (achieved consecutive RT-PCR -). These data indicate that the criterion of ‘≥ 4-fold increase in IgG titer’ might be too stringent to confirm a minority of COVID-19 patients.

### Identification of COVID-19 patients in suspects with negative RT-PCR results

To evaluate the potentiality of the serological test in COVID-19 diagnosis, 52 COVID-19 suspects admitted to Wanzhou People’s Hospital (Chongqing, China) who had respiratory symptoms or abnormal pulmonary imaging, but with negative RT-PCR in at least 2 sequential tests were enrolled. Four of the 52 patients showed positive IgG (3/4) or IgM (3/4) in their first samples (Fig. 4). Patients 3 showed an over 4-fold increase in IgG at the latter time point. Interestingly, RT-PCR test of patient 3’s sample became positive one day between the two sampling, confirming the infection of SARS-CoV-2. An increase in the titer of IgM was observed in the 3 sequential samples from patient 1 (< 2-folds). Patient 2 was positive in both the IgG and IgM. Patient 4 had high IgG and IgM titers, more than 100-fold and 10-fold higher respectively, than the cut-off values at both time points. Although the latter 3 patients did not show a seroconversion or a > 4-fold change of IgG titers between sequential samples, we still support a COVID-19 diagnosis for them.

**Fig. 4.**
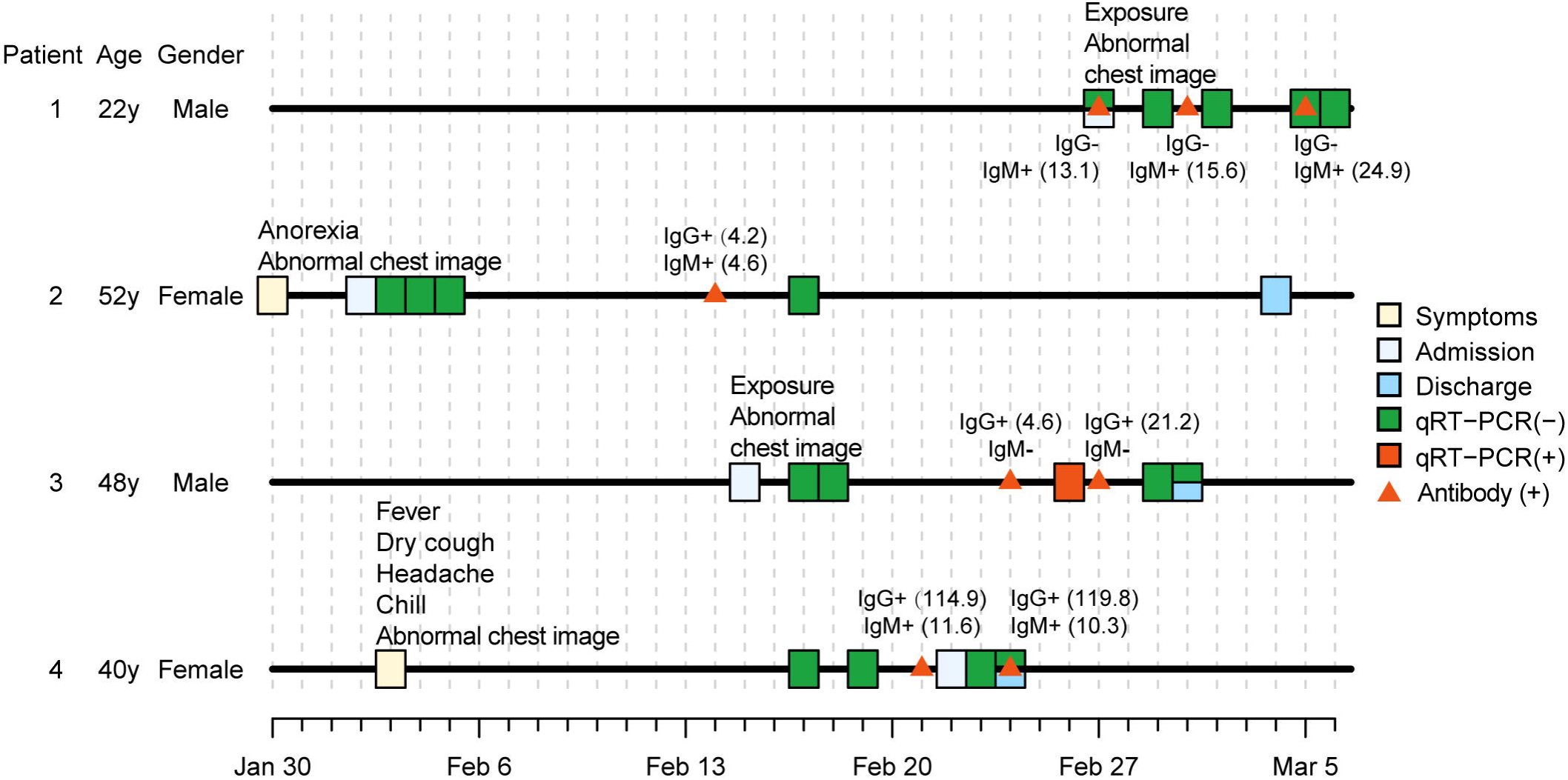
Identification of COVID-19 in suspects. Fifty-two COVID-19 suspects, who failed to be confirmed in successive efforts of sampling and qRT-PCRs, were tested for antibodies. Four of the suspects can be confirmed by the serological test. Numbers in the parentheses are the values of signal/cut-off (S/Co) from the antibody tests, which indicate the titer of the antibodies. S/Co ≥ 1 was determined as positive.

**Fig. 5.**
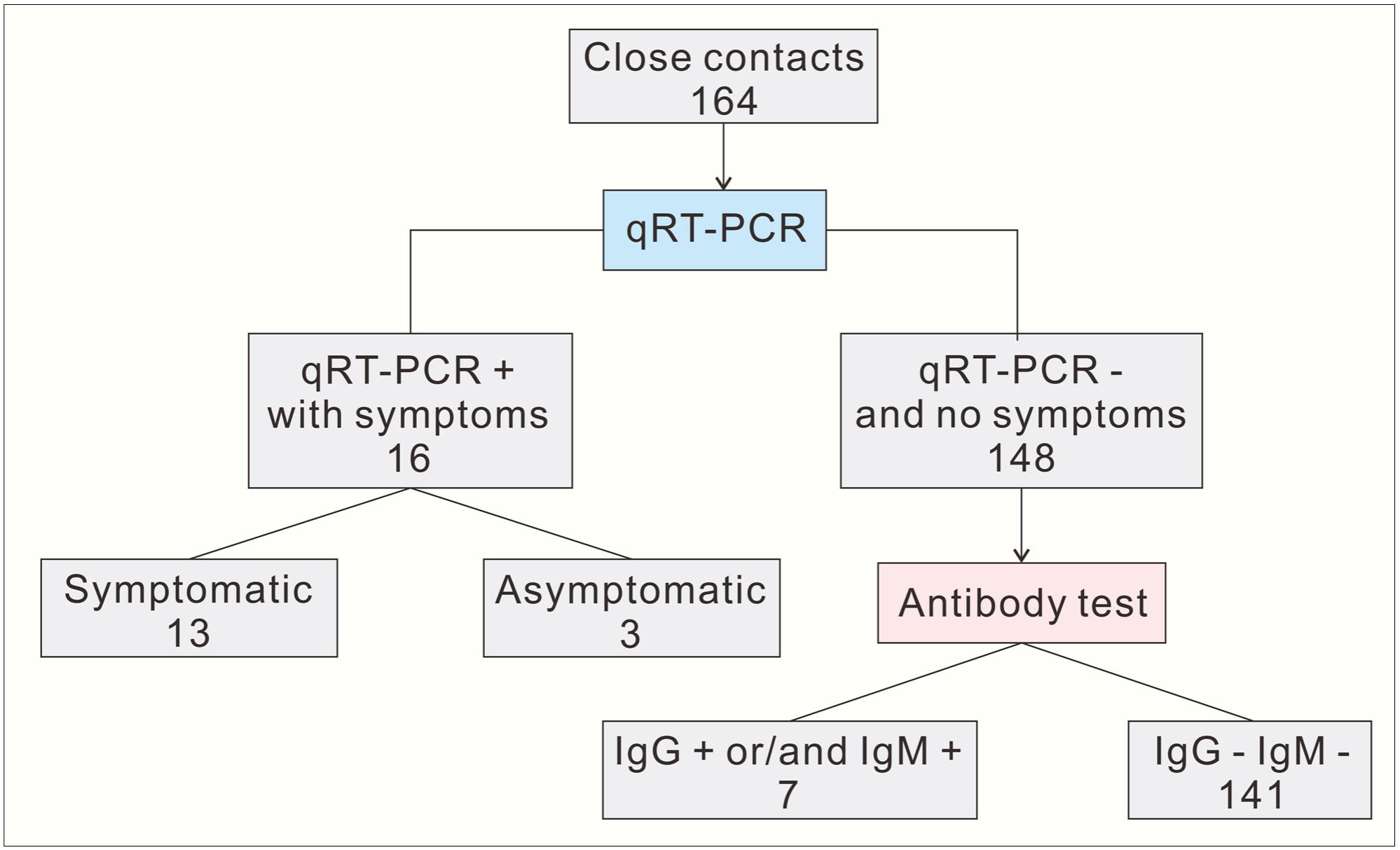
Identification of asymptomatic infection in close contacts by antibody test.

### Identification of asymptomatic infection in close contacts

We next used the serological test in a cluster of 164 close contacts identified by Chongqing CDC. A couple travelled back from Wuhan city, who were confirmed to be SARS-CoV-2 infection on Feb 4, 2020, were deemed as the 1^st^ generation patients of this network. All other cases in this cohort were closely contacted (either directly or indirectly) with this couple during Jan 20 to Feb 6, 2020. A total of 16 out of 164 cases were confirmed by RT-PCR during Jan 31 to Feb 9, 2020, with 3 cases reporting no symptoms. The other 148 cases were no symptoms and negative in RT-PCR tests. On March 1, 2020, serum samples were collected from these 164 cases for antibody tests. The 16 RT-PCR confirmed cases were positive in IgG or/and IgM. Strikingly, 7 of the 148 cases who were excluded previously by negative nucleic acid results also showed positive results in IgG or/and IgM, indicating that 4.3% (7/162) of close contacts were missed by nucleic acid test. In addition, about 6.1% (10/164) of this cohort were asymptomatic infection.

## Discussion

### Proposals for the application of SARS-CoV-2 antibody test in clinical practice

The features of the antibody responses against SARS-CoV-2 give us hints for the application of the serological test in the diagnosis of COVID-19. Firstly, almost all confirmed patients achieve IgG or IgM seroconversion within 20 days after symptoms onset as evidenced by both the cross-section analysis and the follow-up study. This finding indicates that SARS-CoV-2 infection can be ruled out if antibody against SARS-CoV-2 is still undetectable after 20 days of symptoms onset, or after 23 days from exposure (20 days plus a median incubation of 3-day [9]). A mother and her daughter did not achieve a seroconversion of either IgG or IgM during the hospitalization, emphasizing the necessity of a combination of antibody and RT-PCR tests. Secondly, there is no general rule for the chronological order of IgM and IgG seroconversion for a specific patient, which resembles the conditions in SARS and MERS [2, 11, 12]. This supports the detection of both the IgG and IgM simultaneously rather than the single antibody alone. Thirdly, the confirmation criteria of the serological test for MERS recommended by the WHO can apply to a majority of COVID-19 patients. 82.9% (34/41) of the patients can be diagnosed by the 2 criteria: seroconversion or a ≥ 4-fold increase in the IgG titer in acute (ideally during the first week of illness) and convalescent serum samples [10]. The leading cause for the missing of the patients (12.2% (5/41)) by the criteria is a very early seroconversion (within 7 days or even -4 days of symptoms onset) of IgG in these patients (Fig. 3B, middle panel). This finding highlights the importance of collecting the first sample as early as possible. On the other hand, we argue that for those patients missed the ideal sampling window, a second serological test several days (for example, 3 days) later that confirms the first positive results would be sufficient to confirm the diagnosis, if with symptoms or chest imaging evidence simultaneously.

### Serologic test helps to identify asymptomatic infection

The asymptomatic infection poses a special challenge in the prevention of COVID-19, since symptoms are usually used as important indicator for COVID-19. Asymptomatic individual with infection will become a transmission source if not be contained and quarantined. We surveyed a cohort of 164 close contacts and identified 4.3% (7/164) patients with occult infection which were missed by symptoms screening and nucleic acid test. Adding those identified by RT-PCR, the percentage of asymptomatic infection was as high as 6.1% in this cohort. Thus, it is necessary to identify and isolate close contacts even if they showed no symptoms and with negative RT-PCR results. A limitation of the survey is that only one serum sample for each case was obtained. It is better to confirm the test results by second sampling if possible.

In summary, we depict a relatively complete prospect of acute antibody response against SARS-CoV-2 in COVID-19 patients. We believe the antibody test would be of great help in the diagnosis of COVID-19 and the epidemic survey of SARS-CoV-2 infection.

## Data Availability

Data referred to in the manuscript are available.

## Notes

## Acknowledgments

We thank Chengyong Yang, Law Yuen Kwan and Ju Cao for critical reviewing of the manuscript.

## Financial support

This work was supported by the Emergency Project from the Science & Technology Commission of Chongqing; The Major National S&T program grant (2017ZX10202203 and 2017ZX10302201) from Science & Technology Commission of China.

## Potential Conflicts of Interest

Conflicts that the editors consider relevant to the content of the manuscript have been disclosed.

## Notes

### Competing Interest Statement

The authors have declared no competing interest.

